# Demographic and clinical features associated with in-hospital mortality in Egyptian COVID-19 patients: A retrospective cohort study

**DOI:** 10.1101/2021.03.22.21253577

**Authors:** Noha Asem, Mohamed Hassany, Khaled Taema, Hossam Masoud, Gehan Elassal, Ehab Kamal, Wagdy Amin, Akram Abdelbary, Amin Abdel Baki, Samy Zaky, Ahmad Abdalmohsen, Hamdy Ibrahim, Mohamed Elnady, Ahmed Mohamed, Ehab Atteia, Hala Zaid

## Abstract

**Introduction:** Since the worldwide emergence of the COVID-19, several protocols were used by different healthcare organisations. We evaluated in this study the demographic and clinical characteristics of COVID-19 disease in Egyptian population with special consideration for its mortality predictors.

**Methodology:** 8162 participants (mean age 48.7±17.3 years,54.5% males) with RT-PCR positive COVID-19 were included. The electronic medical records were reviewed for demographic, clinical, laboratory, and radiologic features. The primary outcome was the in-hospital mortality rate.

**Results:** The in-hospital mortality was 11.2%. There was a statistically significant strong association of in-hospital mortality with age >60 years old (OR:4.7; 95% CI 4.1-5.4;p<0.001), diabetes mellitus (OR:4.6; 95% CI 3.99-5.32;p<0.001), hypertension (OR:3.9; 95% CI 3.4-4.5;p<0.001), coronary artery disease (OR:2.7; 95% CI 2.2-3.2;p<0.001), chronic obstructive pulmonary disease (OR:2.1; 95% CI 1.7-2.5;p<0.001), chronic kidney disease (OR:4.8; 95% CI 3.9-5.9;p<0.001), malignancy (OR:3.7; 95% CI 2.3-5.75;p<0.001), neutrophil-lymphocyte ratio >3.1 (OR:6.4; 95% CI 4.4-9.5;P< 0.001), and ground glass opacities (GGOs) in CT chest (OR:3.5; 95% CI 2.84-4.4;P<0.001), respectively. There was a statistically significant moderate association of in-hospital mortality with male gender (OR:1.6; 95% CI 1.38-1.83;p<0.001) and smoking (OR:1.6; 95% CI 1.3-1.9;p<0.001). GGOs was reported as the most common CT finding (occurred in 73.1% of the study participants).

**Conclusions:** This multicenter, retrospective study ascertained the higher in-hospital mortality rate in Egyptian COVID-19 patients with different comorbidities.

## INTRODUCTION

A pneumonia outbreak caused by a novel coronavirus, the severe acute respiratory syndrome coronavirus-2 (SARS-CoV-2), has been identified at the end of 2019 in Wuhan city, China. The pneumonia outbreak spread rapidly crossing international boundaries and was declared as a global pandemic by the WHO on March 11^th^, 2020 [1,2]. The disease is designated COVID-19, which stands for coronavirus disease 2019 [3].

Acute COVID-19 pneumonia induced acute respiratory distress syndrome (ARDS) is a potential significant concern for morbidity and mortality, besides other induced life-threatening complications that include, but are not limited to, arrhythmias, acute cardiac injury, and shock. Critical illness mainly occurs in elderly individuals, especially with underlying comorbidities as diabetes mellitus (DM), hypertension, coronary artery disease (CAD), malignancy, chronic obstructive pulmonary disease (COPD), and/or chronic kidney disease (CKD) [4]. Laboratory parameters associated with severe disease and related to disease progression include ferritin and D-Dimer [5].

On 15/02/2020, Egypt declared the first reported COVID-19 patient [6] which increased to a range of 710–5241 patients by March 31^st^ [7]. In February 2020, the Egyptian Ministry of Health designated a scientific committee for steering the national clinical COVID-19 management protocol. Due to lack of evidence for the effectiveness of any antiviral therapy in the management of COVID-19, resource allocation to repurpose the utilization of antiviral and non-antiviral medications approved for the treatment of other diseases was a potential mitigation measure for the current pandemic [8]. It is reported that both chloroquine and hydroxychloroquine (potent inhibitors of DNA and RNA polymerase reactions) inhibit SARS-CoV-2 in vitro [9,10]. These medications are authorized as antimalarials and used in the management of autoimmune disorders, including systemic lupus erythematosus and rheumatoid arthritis. Considering its’ in vitro activity [9,10], safety for inpatient use [11], availability, and effectiveness in some pilot non-randomized studies [12–14], the scientific committee advocated the use of hydroxychloroquine as the main antiviral therapy in the Egyptian COVID-19 management protocol.

We intended to investigate the demographic, clinical, laboratory, and radiologic features associated with in-hospital mortality in the Egyptian COVID-19 patient population.

## METHODS

### Study Design

This was an observational, multicenter, retrospective cohort study conducted at 17 COVID-19 quarantine hospitals in Egypt from March 20^th^ till May 30^th^, 2020. The study design was approved by the research ethics committee and the institutional review board of the Egyptian Ministry of Health and Population. The informed consent for data collection was obtained from patients or first degree relatives, the study procedures were carried out following the Code of Ethics of the World Medical Association (Declaration of Helsinki), all information/images were anonymized and coded, and the privacy and confidentiality rights of the study participants were observed throughout the study.

### Study Participants

Study participants were patients with symptoms suggestive of COVID-19 referred for admission. They were subjected to history taking and data collection for age, gender, smoking, pregnancy, comorbidities especially COPD, rheumatic heart disease, DM, hypertension, CAD, chronic liver disease, CKD, autoimmune disease and malignancy, and nasopharyngeal or oropharyngeal swab for reverse transcription-polymerase chain reaction (RT-PCR) of SARS-CoV-2. Screened participants were enrolled if they were admitted with COVID-19 disease, confirmed by RT-PCR positive nasopharyngeal or oropharyngeal swab, and excluded if they were RT-PCR negative COVID-19 disease.

### Study Procedures

A clinical COVID-19 management protocol was prepared by the Egyptian Ministry of Health COVID-19 scientific steering committee. An educational task force was established to update the healthcare providers on the COVID-19 management protocol. Several hospitals were designated for quarantine of patients with COVID-19. All study participants were followed retrospectively from the admission date to the discharge date. Nasopharyngeal or oropharyngeal swab for RT-PCR of SARS-CoV-2 was repeated every 72 hours after clinical remission of COVID-19 symptoms. The discharge criteria were the absence of the COVID-19 symptoms for at least 3 days, improvement in the radiologic findings, and two consecutive negative nasopharyngeal or oropharyngeal swabs for RT-PCR of SARS-CoV-2 obtained at least 48 hours apart. Epidemiological, demographic, clinical, laboratory, radiologic, treatment, and outcome data were extracted from the study participants’ standardized electronic medical records designated for COVID-19 quarantine hospitals.

### End Points

The primary outcome was the in-hospital mortality rate while the secondary outcome was the negative RT-PCR conversion rate and the time to negative RT-PCR conversion.

### Statistical Analysis

Retrospective data from the study participants’ standardized electronic medical records were collected and coded, and the data were analyzed using the statistical package for the social sciences software (SPSS version 25). A quantitative (continuous) variable was considered parametrically (normally) distributed if the z-value of skewness and kurtosis was between − 1.96 and +1.96 [15] and the Shapiro-Wilk’s test had a p-value > 0.05 [16]. We found that our quantitative (continuous) variables were parametrically (normally) distributed. Accordingly, quantitative (continuous) data were expressed as means and standard deviations. Parametrically distributed quantitative (continuous) variables were compared with the Independent two-tailed t-test. Qualitative (categorical) data were expressed as proportions. Qualitative (categorical) variables were compared with Chi-Square Test (*x*^*2*^). The probability of in-hospital mortality was expressed as odds ratio (OR). The confidence interval was set to 95% (95 % CI) and the margin of error accepted was set to 5%. Any comparison considered statistically significant was at P-value ≤ 0.05.

## RESULTS

### Study Participants and Procedures

We reviewed the standardized electronic medical records of 8162 patients with RT-PCR positive COVID-19 disease from 17 hospitals in one country from March 20^th^ till May 30^th^, 2020. The study group was disproportionate with regards to the demographic data, comorbidities, disease severity, and baseline laboratory findings (Table 1). The key sociodemographic feature of the enrolled participants was slight male predominance (Mean age 48.7 ± 17.3 years, 54.5% males, 45.5% females). All enrolled participants completed the study and there were no withdrawals. The main presenting clinical features of the study participants were fever (69.8%; 95% CI 68.8% − 70.8%) and cough (67.5%; 95% CI 66.5% - 68.5%), respectively (Figure 1). The most common CT finding was the ground glass opacities (GGOs) which were reported in 5962 study participants (73.1%; 95% CI 72.1% - 74%). There was septal thickening in 2384 study participants (29.2%; 95% CI 28.2% - 30.2%), consolidation in 2884 study participants (35.3%; 34.3% - 36.4%), lymphadenopathy in 1470 study participants (18%; 95% CI 17.2% - 18.8%), pleural effusion in 653 study participants (8%; 95% CI 7.4% - 8.6%), peripheral subpleural involvement in 4163 study participants (51%; 95% CI 49.9% - 52.1%), and multilobar involvement in 4679 study participants (57.3%; 95% CI 56.3% - 58.4%), respectively. Pneumonia was the most common complication in our cohort, reported in 1425 study participants (17.5%; 95% CI 16.6% - 18.3%). There was acute respiratory distress syndrome (ARDS) in 996 study participants (12.2%; 95% CI 11.5% - 12.9%), cardiac arrhythmias in 112 study participants (1.4%; 95% CI 1.1% – 1.6%), myocarditis in 191 study participants (2.3%; 95% CI 2% - 2.7%), disseminated intravascular coagulopathy in 320 study participants [3.9%; 95% CI 3.5% - 4.3%), bleeding events in 12 study participants (0.15%; 95% CI 0.06% - 0.23%), acute kidney injury in 350 study participants (4.29%; 95% CI 3.9% - 4.7%), acute liver injury in 204 study participants (2.5%; 95% CI 2.2% - 2.8%), and septic shock in 1098 study participants (13.5%; 95% CI 12.7% - 14.2%), respectively.

**Table 1:**
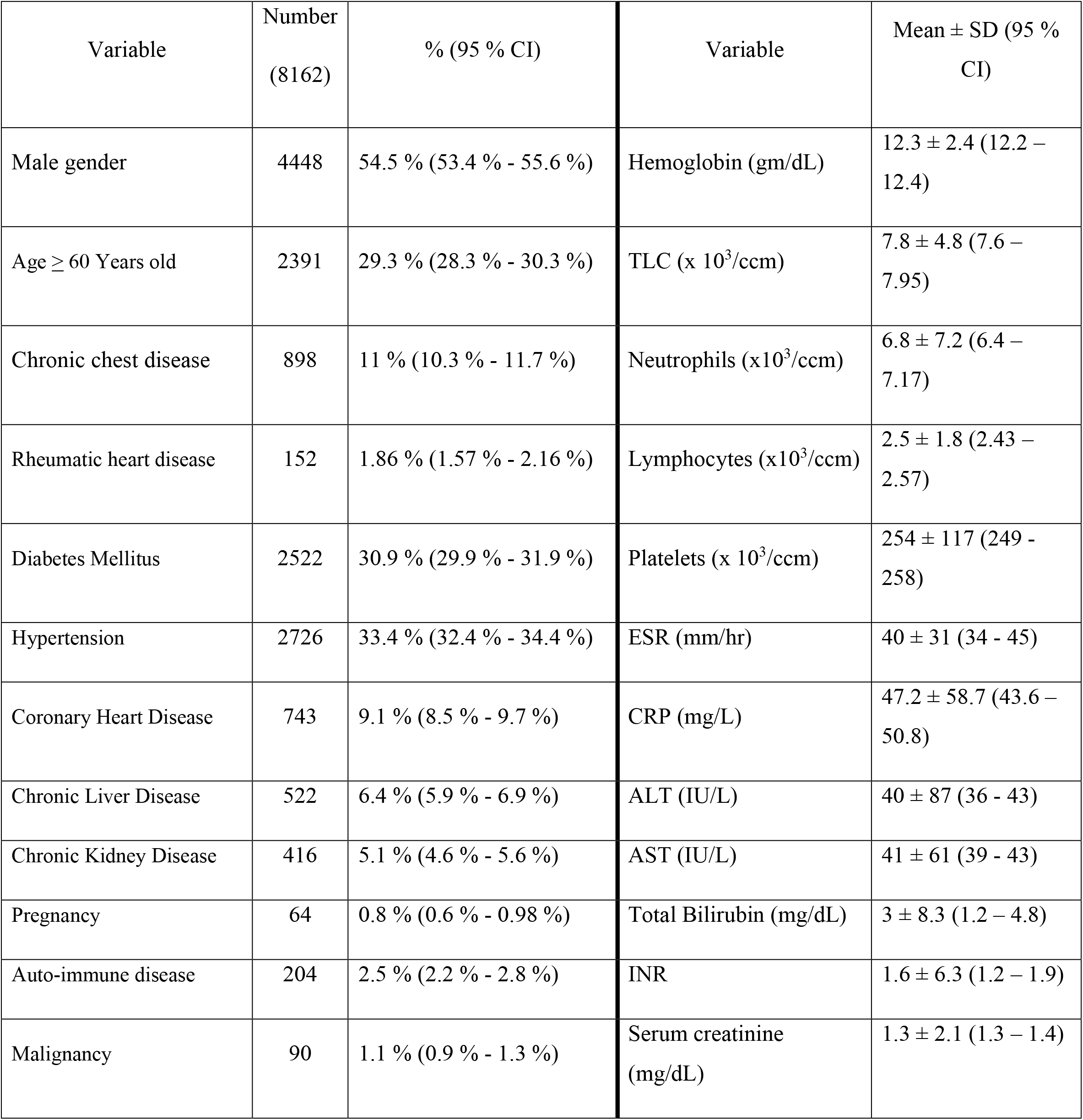
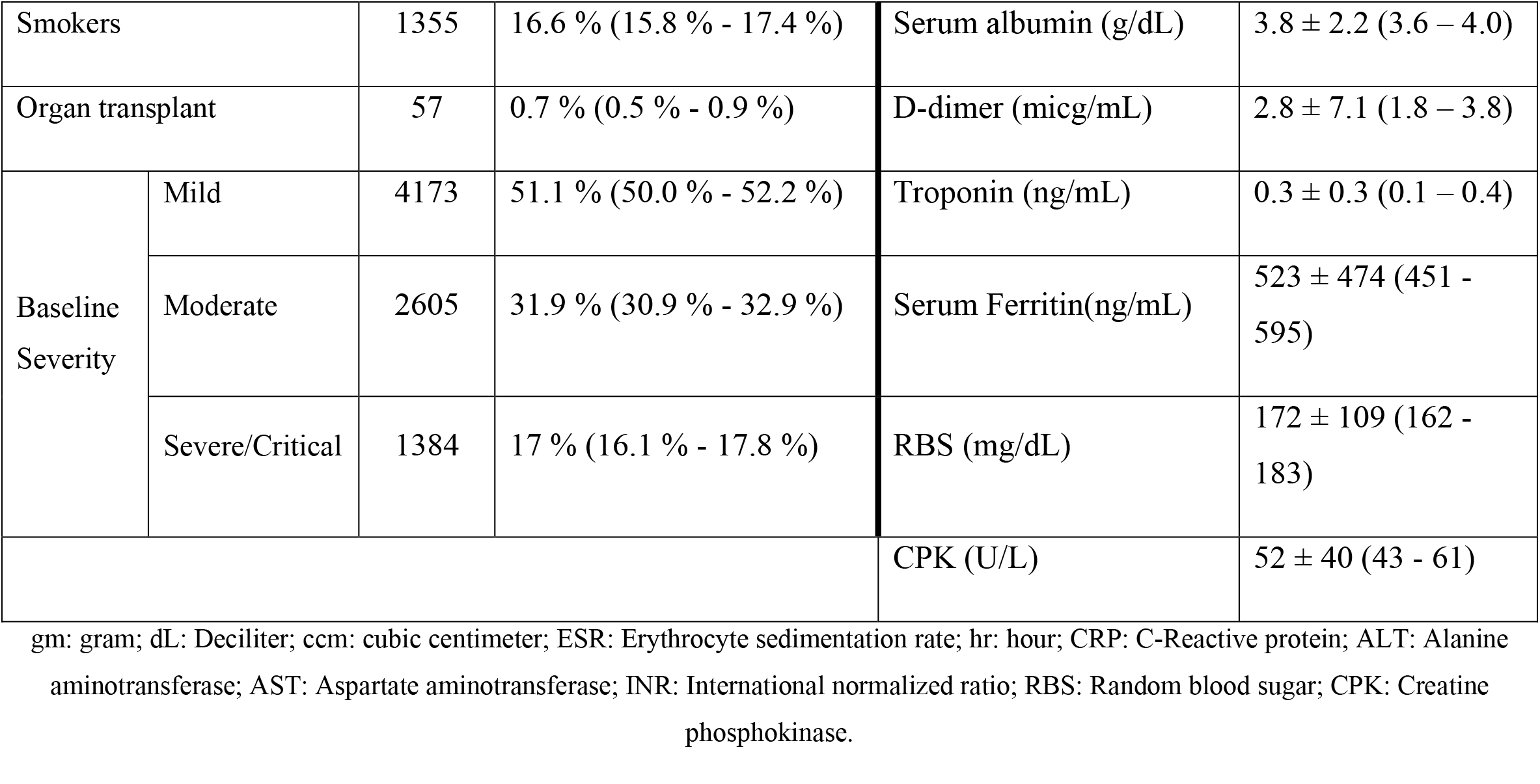
The demographic data, comorbidities, disease severity, and baseline laboratory findings of the study population

| Variable | Number<br>(8162) | % (95 % CI) | Variable | Mean ± SD (95 %<br>CI) |
| --- | --- | --- | --- | --- |
| Male gender | 4448 | 54.5 % (53.4 % - 55.6 %) | Hemoglobin (gm/dL) | 12.3 ± 2.4 (12.2 – 12.4) |
| Age ≥ 60 Years old | 2391 | 29.3 % (28.3 % - 30.3 %) | TLC (x 10<sup>3</sup>/ccm) | 7.8 ± 4.8 (7.6 – 7.95) |
| Chronic chest disease | 898 | 11 % (10.3 % - 11.7 %) | Neutrophils (x10<sup>3</sup>/ccm) | 6.8 ± 7.2 (6.4 – 7.17) |
| Rheumatic heart disease | 152 | 1.86 % (1.57 % - 2.16 %) | Lymphocytes (x10<sup>3</sup>/ccm) | 2.5 ± 1.8 (2.43 – 2.57) |
| Diabetes Mellitus | 2522 | 30.9 % (29.9 % - 31.9 %) | Platelets (x 10<sup>3</sup>/ccm) | 254 ± 117 (249 - 258) |
| Hypertension | 2726 | 33.4 % (32.4 % - 34.4 %) | ESR (mm/hr) | 40 ± 31 (34 - 45) |
| Coronary Heart Disease | 743 | 9.1 % (8.5 % - 9.7 %) | CRP (mg/L) | 47.2 ± 58.7 (43.6 – 50.8) |
| Chronic Liver Disease | 522 | 6.4 % (5.9 % - 6.9 %) | ALT (IU/L) | 40 ± 87 (36 - 43) |
| Chronic Kidney Disease | 416 | 5.1 % (4.6 % - 5.6 %) | AST (IU/L) | 41 ± 61 (39 - 43) |
| Pregnancy | 64 | 0.8 % (0.6 % - 0.98 %) | Total Bilirubin (mg/dL) | 3 ± 8.3 (1.2 – 4.8) |
| Auto-immune disease | 204 | 2.5 % (2.2 % - 2.8 %) | INR | 1.6 ± 6.3 (1.2 – 1.9) |
| Malignancy | 90 | 1.1 % (0.9 % - 1.3 %) | Serum creatinine<br>(mg/dL) | 1.3 ± 2.1 (1.3 – 1.4) |

|  |  |  |  |  |  |
| --- | --- | --- | --- | --- | --- |
| Smokers |  | 1355 | 16.6 % (15.8 % - 17.4 %) | Serum albumin (g/dL) | 3.8 ± 2.2 (3.6 – 4.0) |
| Organ transplant |  | 57 | 0.7 % (0.5 % - 0.9 %) | D-dimer (micg/mL) | 2.8 ± 7.1 (1.8 – 3.8) |
| Baseline Severity | Mild | 4173 | 51.1 % (50.0 % - 52.2 %) | Troponin (ng/mL) | 0.3 ± 0.3 (0.1 – 0.4) |
|  | Moderate | 2605 | 31.9 % (30.9 % - 32.9 %) | Serum Ferritin(ng/mL) | 523 ± 474 (451 - 595) |
|  | Severe/Critical | 1384 | 17 % (16.1 % - 17.8 %) | RBS (mg/dL) | 172 ± 109 (162 - 183) |
|  |  |  |  | CPK (U/L) | 52 ± 40 (43 - 61) |

gm: gram; dL: Deciliter; ccm: cubic centimeter; ESR: Erythrocyte sedimentation rate; hr: hour; CRP: C-Reactive protein; ALT: Alanine aminotransferase; AST: Aspartate aminotransferase; INR: International normalized ratio; RBS: Random blood sugar; CPK: Creatine phosphokinase.

**Figure 1:**
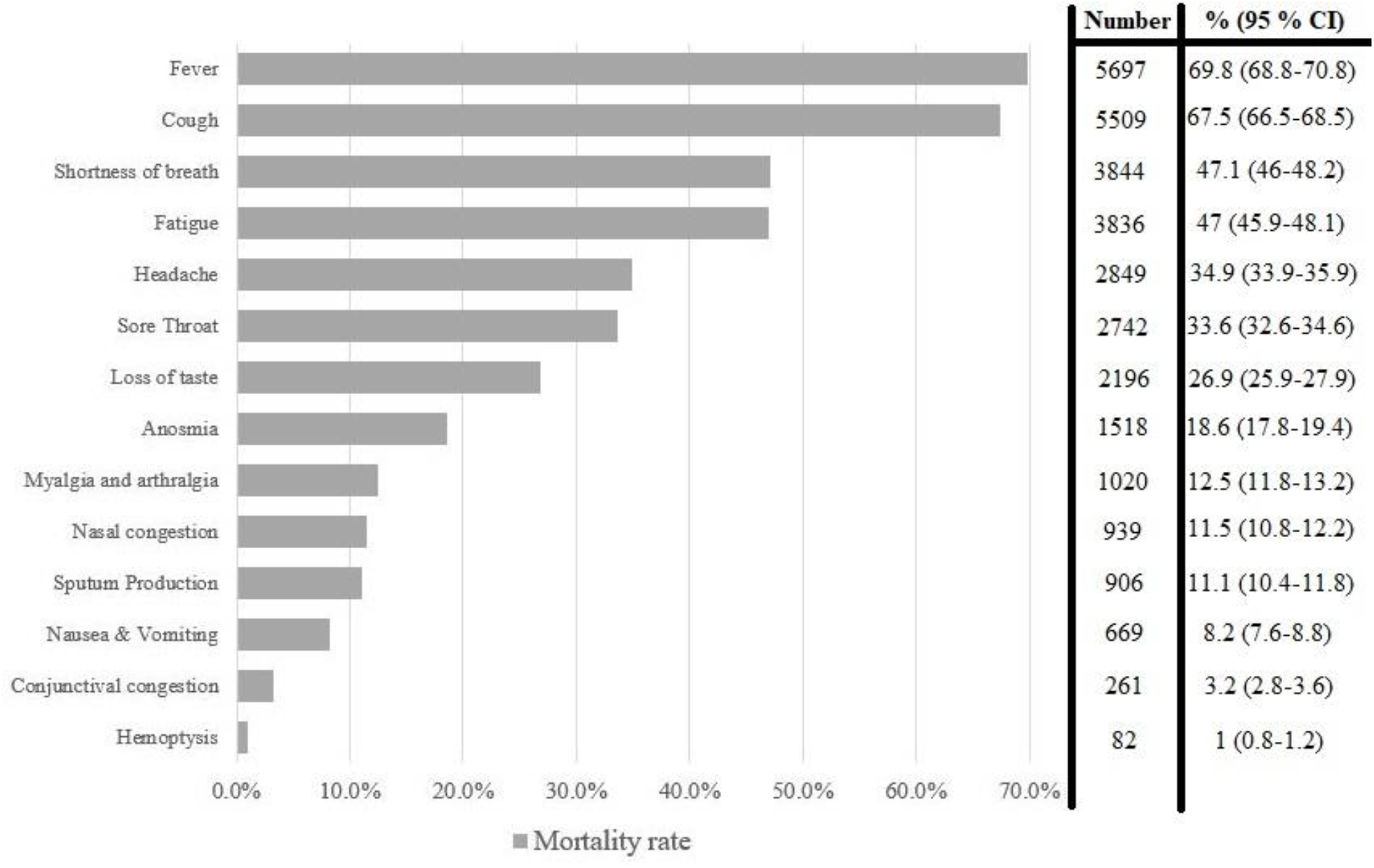
The clinical presentation of the study population

### Clinical, Laboratory, and Radiologic Features associated with In-Hospital Mortality

A total of 7247 study participants survived their hospital course, while 915 study participants died with an in-hospital mortality rate of 11.2% (95% CI 10.5% - 11.9%). The highest mortality rate was in the severe/critical subgroup (646 study participants) (46.7%; 95% CI 44.0% - 49.3%) compared to the mild subgroup (68 study participants) (1.6%; 95% CI 1.2% - 2.0%) and the moderate subgroup (201 study participants) (7.7%; 95% CI 6.7% - 8.7%), respectively (P< 0.001). The in-hospital mortality was 23.5% in patients >60 years old compared to 6.1% in those <65 years old (*P* < 0.001). The in-hospital mortality was reported in 8.3% of patients with no comorbidities (511/6182) compared to 16.3% (159/977), 21% (145/690), and 31.9% (100/313) of patients with one, two, and three or more comorbid conditions, respectively (P < 0.001) (Figure 2).

**Figure 2:**
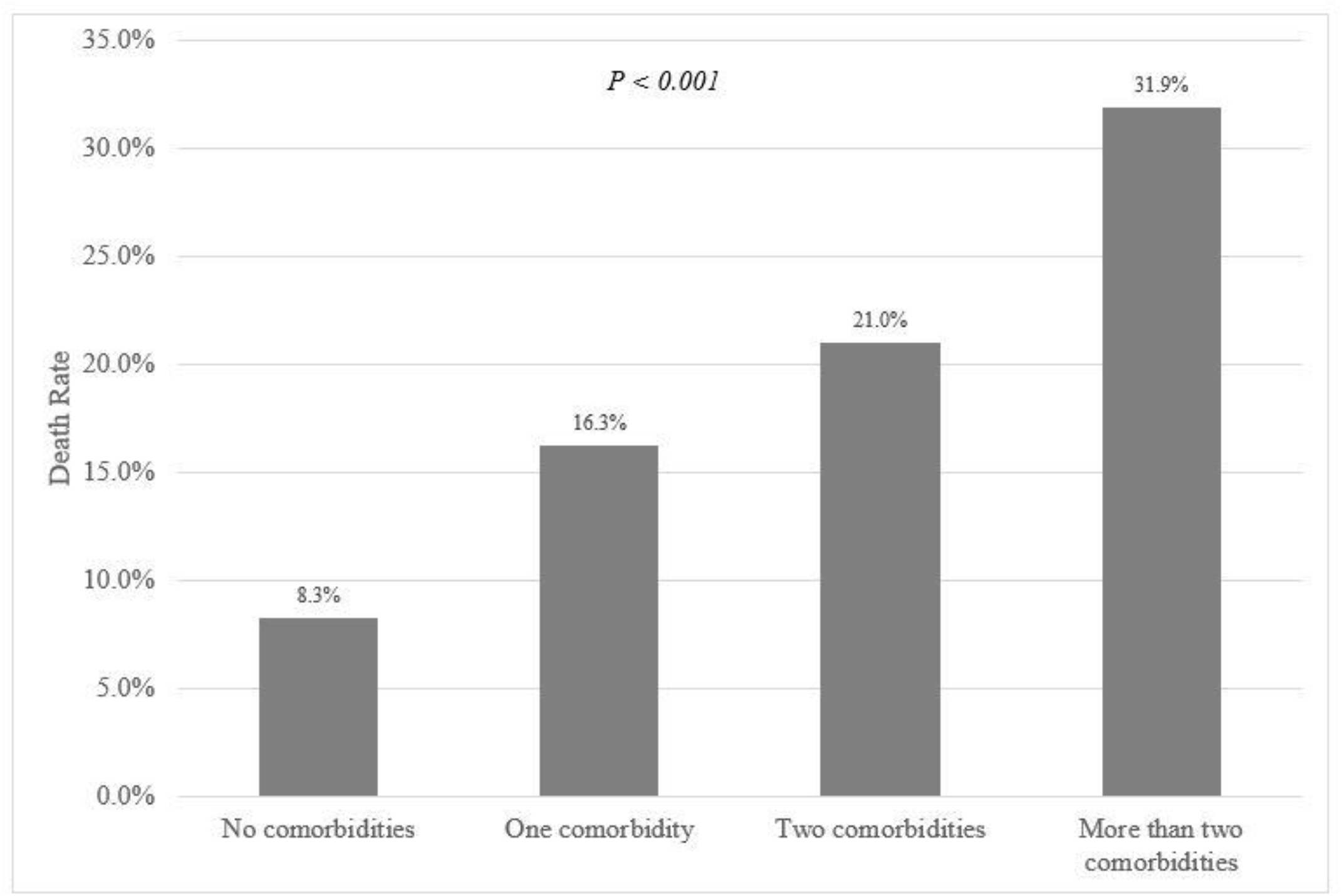
The mortality rate with the different categories of comorbid conditions

There was a statistically significant strong association of in-hospital mortality with age >60 years old (OR: 4.7; 95% CI 4.1-5.4), DM (OR: 4.6; 95% CI 3.99-5.32), hypertension (OR: 3.9; 95% CI 3.4-4.5), CAD (OR: 2.7; 95% CI 2.2-3.2), COPD (OR: 2.1; 95% CI 1.7-2.5), chronic liver disease (OR: 2.5; 95% CI 2.04-3.2), CKD (OR: 4.8; 95% CI 3.9-5.9), and malignancy (OR: 3.7; 95% CI 2.3-5.75), respectively (P< 0.001). There was a statistically significant moderate association of in-hospital mortality with male gender (OR: 1.6; 95% CI 1.38-1.83) and smoking (OR: 1.6; 95% CI 1.3-1.9), respectively (P< 0.001) (Table 2).

**Table 2:** Clinical predictors of mortality

| Variable | Number of patients | Number of deaths | Mortality rate (95 % CI) | OR (95 % CI) | P value |
| --- | --- | --- | --- | --- | --- |
| Male gender | 4448 | 589 | 13.2% (12.3%-14.2%) | 1.6 (1.38-1.83) | < 0.001 |
| 60 or more years old | 2391 | 561 | 23.5% (21.8%-25.2%) | 4.7 (4.1-5.4) | < 0.001 |
| Pregnancy | 64 | 3 | 4.7% (-0.5%-9.9%) | 0.39 (0.12-1.24) | 0.061 |
| Chronic obstructive pulmonary disease | 898 | 171 | 19.0% (16.5%-21.6%) | 2.1 (1.7-2.5) | < 0.001 |
| Diabetes Mellitus | 2522 | 575 | 22.8% (21.2%-24.4%) | 4.6 (3.99-5.3) | < 0.001 |
| Hypertension | 2726 | 569 | 20.9% (19.3%-22.4%) | 3.9 (3.4-4.5) | < 0.001 |
| Coronary heart disease | 743 | 170 | 22.9% (19.9%-25.9%) | 2.7 (2.2-3.2) | < 0.001 |
| Chronic liver disease | 522 | 119 | 22.8% (19.2%-26.4%) | 2.5 (2.04-3.2) | < 0.001 |
| History of rheumatic heart disease | 152 | 32 | 21.1% (14.6%-27.5%) | 2.2 (1.5-3.2) | < 0.001 |
| Chronic kidney disease | 416 | 144 | 34.6% (30.0%-39.2%) | 4.8 (3.9-5.9) | < 0.001 |
| Malignancy | 90 | 28 | 31.1% (21.5%-40.7%) | 3.7 (2.3-5.8) | < 0.001 |
| Smoking | 1355 | 208 | 15.4% (13.4%-17.3%) | 1.6 (1.3-1.9) | < 0.001 |
| Organ transplant recipient | 57 | 3 | 5.3% (-0.5%-11.1%) | 0.44 (0.14-1.4) | 0.43 |

Apart from the occurrence of bleeding, the presence of any of the complications, or the presence of any of the CT findings was associated with a significantly higher risk for mortality (Figures 3 and 4).

**Figure 3:**
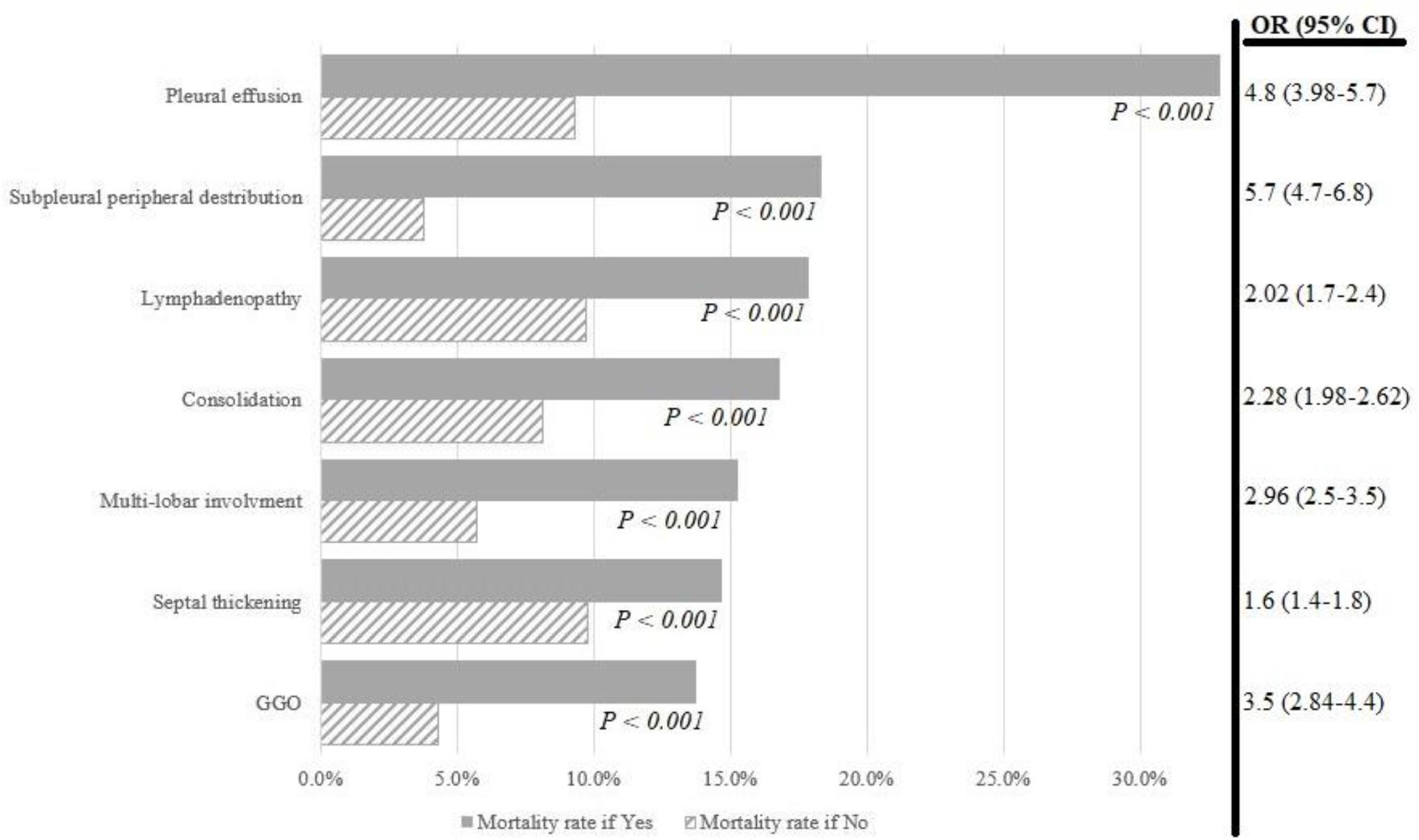
The mortality rate when any of the CT findings is present (Yes) compared to its absence (No)

**Figure 4:**
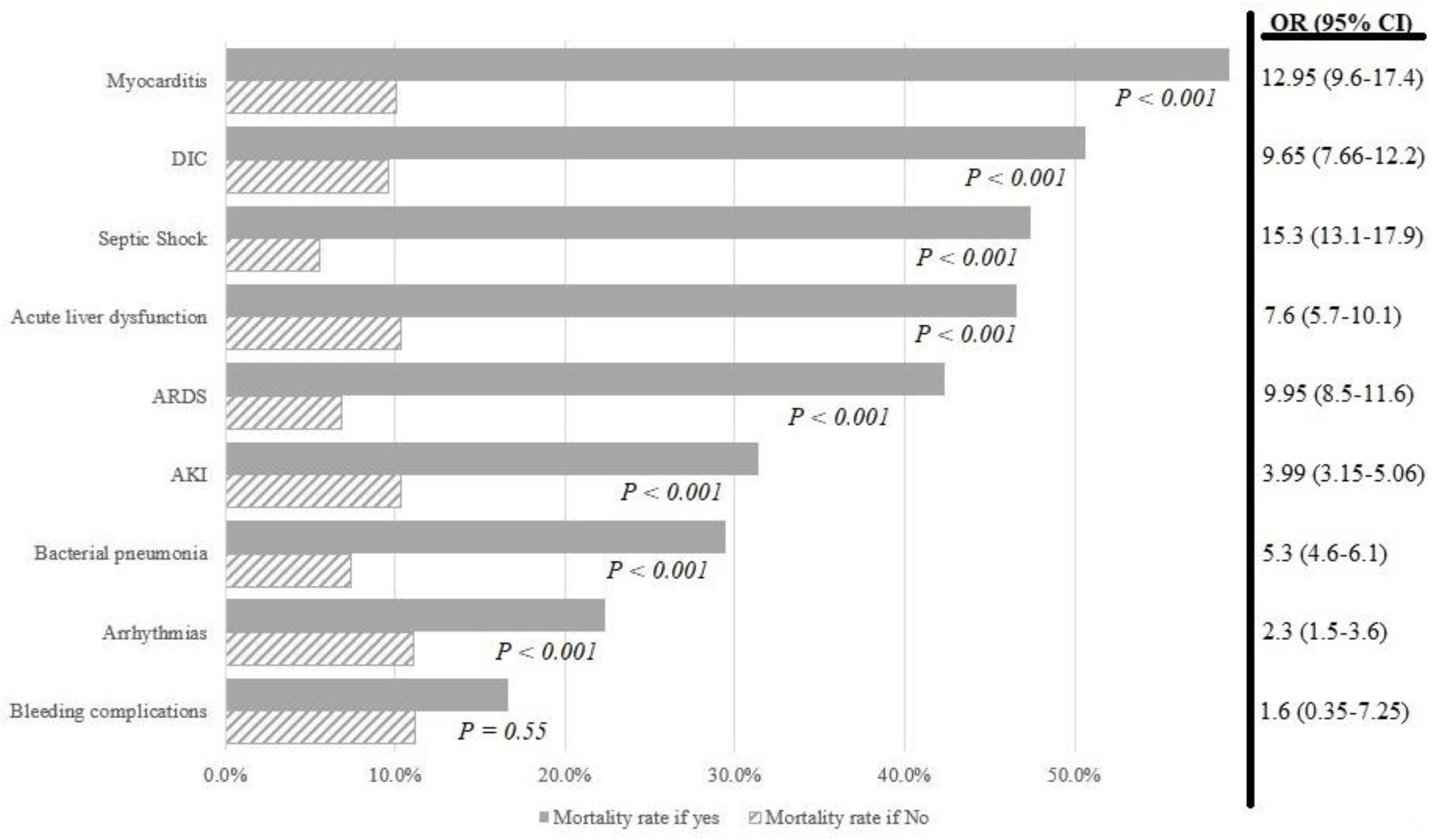
The mortality rate with the different reported complications

The lower serum hemoglobin level, higher total leucocytic count, lower lymphocytic count, higher alanine transaminase (ALT) and aspartate transaminase (AST), higher C-reactive protein (CRP), higher serum creatinine and ferritin levels, lower serum albumin, and higher admission random blood sugar were shown to be significantly associated with higher in-hospital mortality (Table 3).

**Table 3:** The baseline laboratory findings in survivors and non-survivors

| Variable | Non-Survivors | Survivors | P value |
| --- | --- | --- | --- |
| | Mean ± SD (95 % CI) | Mean ± SD (95 % CI) | |
| Hemoglobin (gm/dL) | 11.8 ± 2.2 (11.6-11.9) | 12.4 ± 2.5 (12.3-12.5) | < 0.001 |
| TLC (x 10<sup>3</sup>/ccm) | 11.6 ± 6.4 (11.0-12.2) | 7.1 ± 4.1 (6.9-7.3) | < 0.001 |
| Neutrophils (x 10<sup>3</sup>/ccm) | 10.1 ± 6.7 (9.2-10.97) | 6.2 ± 7.1 (5.8-6.6) | < 0.001 |
| Lymphocytes (x 10<sup>3</sup>/ccm) | 1.5 ± 1.48 (1.35-1.65) | 1.86 ± 1.3 (1.8-1.92) | < 0.001 |
| Platelets (x 10<sup>3</sup>/ccm) | 244 ± 117 (234-255) | 255 ± 117 (251-260) | 0.06 |
| ESR (mm/hr) | 40.8 ± 27.8 (26.7-54.9) | 39.4 ± 31.2 (33.7-45.0) | 0.7 |
| CRP (mg/L) | 74.8 ± 81.9 (62.1-87.4) | 42.1 ± 51.7 (38.6-45.5) | < 0.001 |
| ALT (IU/L) | 55.1 ± 154.5 (39.5-70.7) | 36.2 ± 63.6 (33.3-39.1) | < 0.001 |
| AST (IU/L) | 63 ± 99.0 (53.2-72.8) | 36.4 ± 47.8 (34.2-38.5) | < 0.001 |
| Total Bilirubin (mg/dL) | 1.3 ± 1.1 (0.6-2.0) | 3.2 ± 8.8 (1.2-5.2) | 0.5 |
| INR | 2.1 ± 8.7 (0.9-3.2) | 1.5 ± 5.6 (1.1-1.8) | 0.19 |
| Serum creatinine (mg/dL) | 2.0 ± 2.5 (1.8-2.3) | 1.2 ± 2.0 (1.1-1.3) | < 0.001 |
| Serum albumin (g/dL) | 3.2 ± 0.6 (3.1-3.3) | 3.9 ± 2.4 (3.7-4.1) | < 0.001 |
| D-dimer (micg/mL) | 5.4 ± 8.8 (2.5-8.2) | 2.2 ± 6.5 (1.2-3.2) | 0.05 |
| Troponin (ng/mL) | 0.2 ± 0.3 (-0.1-0.4) | 0.3 ± 0.3 (0.1-0.5) | 0.28 |
| Serum Ferritin(ng/mL) | 919.1 ± 600.5 (688.2-1149.9) | 450.3 ± 409.5 (382.7-517.9) | 0.001 |
| RBS (mg/dL) | 213.1 ± 143.5 (179.8-246.5) | 163.6 ± 98.4 (153.0-174.1) | 0.007 |
| CPK (U/L) | 56.8 ± 40.5 (34.8-78.9) | 51.2 ± 40.7 (41.8-60.7) | 0.65 |

gm: gram; dL: Deciliter; ccm: cubic centimeter; ESR: Erythrocyte sedimentation rate; hr: hour; CRP: C-Reactive protein;
ALT: Alanine aminotransferase; AST: Aspartate aminotransferase; INR: International normalized ratio; RBS: Random
blood sugar; CPK: Creatine phosphokinase.

Neutrophil lymphocyte ratio (NLR) was available in 1398 study participants in the moderate and severe/critical subgroups only. A total of 192 study participants with NLR >3.1 died (26 %) compared to 34 study participants with NLR <3.1 (5.2 %). There was a statistically significant strong association of in-hospital mortality with NLR >3.1 (OR: 6.4; 95% CI 4.4-9.5) and GGOs in CT chest (OR: 3.5; 95% CI 2.84-4.4), respectively (P< 0.001).

### Negative RT-PCR Conversion Rate and Time to Negative RT-PCR Conversion

The negative RT-PCR conversion rate was 86.2% by the 12^th^ day of treatment and 95.9% after the 18^th^ day of treatment with a peak of 15.2% on the 3^rd^ day of treatment (Figure 5).

**Figure 5:**
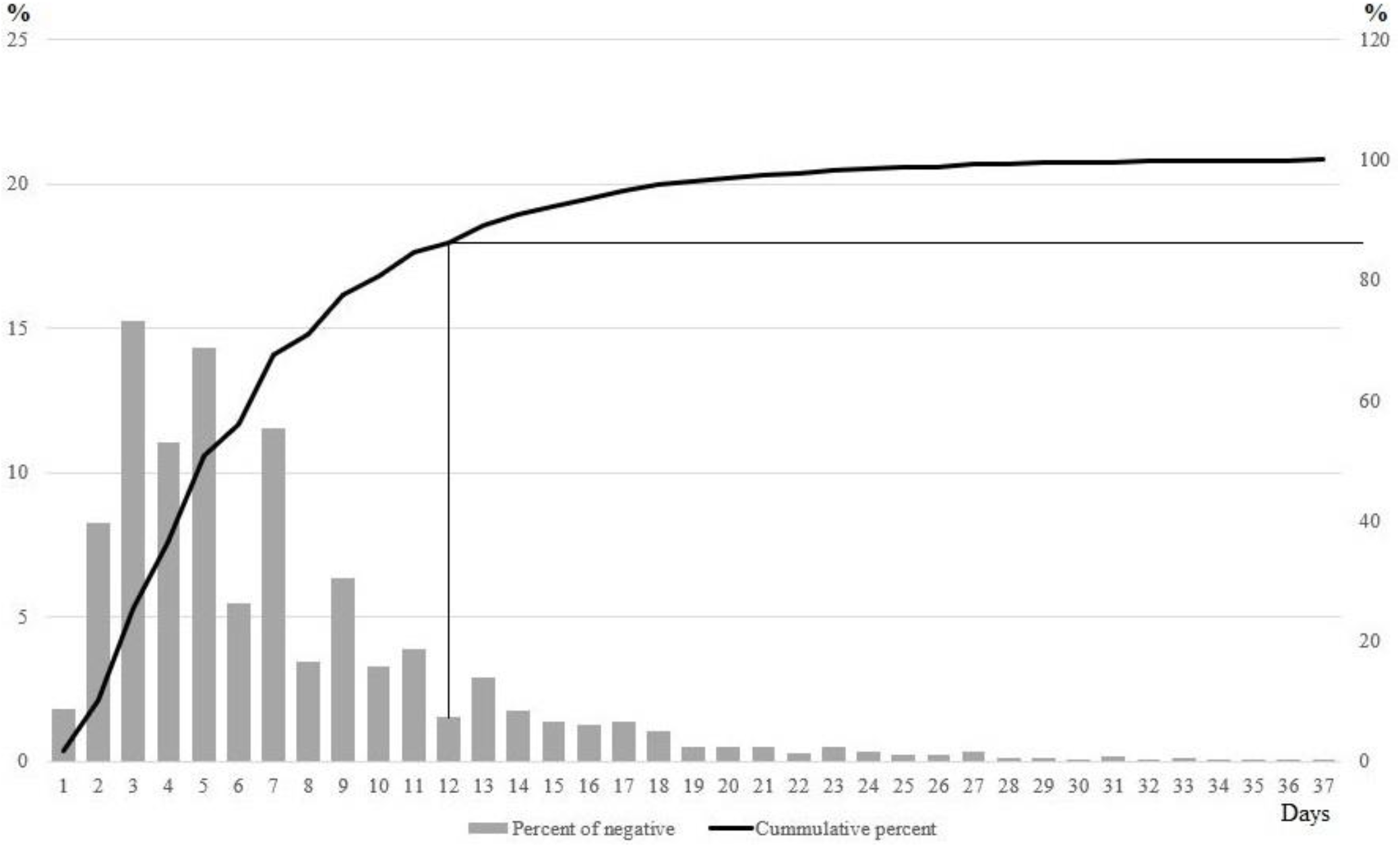
The pattern of turning PCR negative after starting treatment protocol

## DISCUSSION

With the emergence of the first COVID-19 patient in Egypt on 15/02/2020 [6], the Egyptian Ministry of Health and Population designated a scientific committee for steering a national COVID-19 management protocol to admit and treat COVID-19 patients in quarantine hospitals. Considering its’ in vitro activity [9,10], safety for inpatient use [11], availability, and effectiveness in some pilot non-randomized studies [12–14], the scientific committee advocated the use of hydroxychloroquine as the main antiviral therapy in the Egyptian COVID-19 management protocol. In this study, we analyzed data of 8162 patients with RT-PCR positive COVID-19 disease admitted in 17 Egyptian quarantine hospitals from March 20^th^ till May 30^th^, 2020.

Fever and dry cough were the most common presentations of COVID-19 patients [17–19]. Fever and cough were reported in 69.8% and 67.5% of our study participants. In another study [17], fever was reported in 94% of patients and cough in 79% of patients. These symptoms of presentation were not, however, associated with mortality [17].

Our study’s overall in-hospital mortality was 11.2 %. According to the baseline severity of illness identified on presentation to the healthcare facilities, the mortality rates were 1.6%, 7.7%, and 46.7% for mild, moderate, and severe/critical cases, respectively. A retrospective cohort study of 191 hospitalized COVID-19 patients in Wuhan, China from December 29^th^, 2019 till January 31^st^, 2020 reported the death of 54 patients with a mortality rate of 28.3 %. However, they didn’t report the stratification of mortality as per disease severity. In a meta-analysis of 14 studies including 4659 hospitalized COVID-19 patients, the overall mortality rate was 25.5 % [19]. A higher mortality rate of 32.7 % was reported in high-risk severe patients admitted to ICU in Lombardy, Italy. The in-hospital mortality rates published in these studies are comparable to ours.

In addition to the baseline severity of illness identified on hospital admission and the occurrence of complications during hospital course, the clinical and laboratory features associated with in-hospital mortality were age >60 years old, male gender, COPD, DM, hypertension, CAD, chronic liver disease, malignancy, CKD, neutrophilia, lymphopenia, high CRP, ALT and AST levels, and higher admission serum creatinine and ferritin levels. The Odds Ratio reported in our study suggested a strong association of the in-hospital mortality with age >60 years old, DM, hypertension, chronic liver disease, CKD, malignancy, septic shock, myocarditis, ARDS, DIC, acute liver dysfunction, lymphadenopathy, pleural effusion, and CT finding of multilobar involvement.

Several studies revealed higher mortality rates with advancing age [18,20,21]. A study by Juan Berenguer involving 4035 patients in Spain showed a higher mortality rate with increasing age >60 years old (OR: 7.5; 95% CI 6-9.4) compared to (OR: 4.7; 95% CI 4.1-5.4) reported in our study [22]. This might be secondary to comorbidities linked to advancing age and decreased immune response. CKD, malignancy and liver cirrhosis were reported to be strongly associated with mortality in COVID-19 patients with an OR for CKD ranging from 3.6 in many studies to 9.4 in one meta-analysis study, and OR for malignancy and liver cirrhosis of 2.3 to 2.8, respectively [19,22]. In addition to their significant impact on mortality, the higher incidence rates of hypertension (33.4%) and DM (30.9%) add to their importance as measures of association with mortality in COVID-19 patients. Hypertension and DM were reported as the most common comorbidities that impact mortality in many studies [19,22]. In our study, hypertension and DM caused 3.9 and 4.6 folds increase in the odds of death in comparison to other studies [17–19,22]. The previously known endothelial dysfunction reported with hypertension and DM might explain their impact on the severity of the disease and the high mortality rate [23,24]. Similar to other studies, our study reported that the presence of more comorbidities increases the mortality rates apart from the type of comorbidities [22].

Leukocytosis, lymphopenia, high serum CRP, ALT, AST, and ferritin levels reported in our study were associated with higher mortality rates. Similarly, Berenguer et al reported significantly higher mortality with higher neutrophil count and lower lymphocyte count. They reported also higher CRP, ALT, and AST in non-survivors [22]. The association of higher mortality rates with leukocytosis, neutrophilia, high CRP, and high ferritin levels might be secondary to the dysregulated immune response in COVID-19 patients or superadded secondary infections [25]. The higher mortality rate associated with lymphopenia might represent a dysregulated cellular immune response characterizing COVID-19 [26].

Combining both, the higher NLR was associated with higher mortality. We reported an in-hospital mortality rate of 26% in patients with NLR >3.1 compared to 5.2% in patients with NLR <3.1. Similar results were reported in earlier studies [27]. The elevated serum creatinine, ALT, AST, and low albumin levels might explain the higher mortality reported in chronic liver disease and CKD. We reported a non-significant tendency for higher D-dimer levels in non-survivors. Contrary to this, a significant tendency for higher D-dimer levels in non-survivors was reported by many other investigators [17,28] representing the coagulopathy pattern in COVID-19.

The most common CT finding was the GGOs which were reported in 73.1% of patients. Its’ presence showed a more than 3 folds increase in the odds of death in our cohort of study participants. Despite their lower incidence of occurrence, the presence of consolidation, pleural effusion, or lymphadenopathy was associated with a significantly higher mortality rate indicating a complicated hospitalization course. The GGOs were reported to be the most common CT findings, and measure of severity and association with mortality in many other studies [17,18].

Bacterial pneumonia was reported in >17 % of our study participants. Other studies have reported pneumonia in 10.6% to 91.1% of their study participants [18,22]. The incidence rates of acute kidney injury, septic shock, ARDS and other complications reported in our study were comparable to other studies [17]. The incidence rates of cardiac arrhythmias and acute liver injury were 1.4% and 2.5%, respectively, despite using hydroxychloroquine as a standard of care in the Egyptian COVID-19 management protocol. This contradicts the safety concern about the use of hydroxychloroquine in hospitalized COVID-19 patients [29,30].

Our study showed negative RT-PCR conversion rates of 15.2% by the 3^rd^ day of treatment and 80.7% by the 10^th^ day of treatment. Xiaowen et al. revealed similar negative RT-PCR conversion rates of 10.2% by the 7^th^ day of treatment, 62.7% by the 14^th^ day of treatment, and 91.2% by the 21^st^ day of treatment, respectively [31]. They also stated that elderly patients and patients with chest tightness are more likely to have delayed negative RT-PCR conversions [31].

## STRENGTHS AND LIMITATIONS

Our study has strengths. There were on-site visits by the scientific steering committee members to verify compliance with the Egyptian COVID-19 management protocol. The incidence rates of the clinical efficacy outcomes were estimated, information was collected on all relevant potential confounders, recall bias was prevented, and the causal inference was strengthened. On the other hand, the study has limitations. There were inadequate reporting and analysis of our interventions data so, couldn’t specify the pivotal medication which played an essential role in the management of the COVID-19 disease. Updates to the Egyptian COVID-19 management protocol were not applied to the current study. The documentation process for laboratory investigations in the quarantine hospitals was inefficient in the early phase of the study with subsequent missing data for the continuous laboratory variables. Despite the study limitations, we collected complete data for the categorical variables.

## CONCLUSIONS

This multicenter, retrospective cohort study investigated the demographic, clinical, laboratory, and radiologic features associated with in-hospital mortality in the Egyptian COVID-19 patient population and confirmed the higher in-hospital mortality rate in patients with comorbidities. Double-blinded randomized controlled trials are warranted to make an inference about the treatment effect of the different management elements.

## Data Availability

All data in the manuscript are available

